# Smaller Cortical Responses to Balance Perturbations in Parkinson’s Disease with Freezing of Gait

**DOI:** 10.64898/2026.11.03.26367107

**Authors:** Andrew S. Monaghan, Isaiah J. Lachica, Janna Protzak, Stewart A. Factor, James Lucas Mckay, Michael R. Borich, Lena H. Ting, Jasmine L. Mirdamadi

**Author notes:** Corresponding Author Jasmine Mirdamadi Department of Rehabilitation and Movement Science University of Vermont Burlington, VT.

## Abstract

Freezing of gait (FoG) is among the most disabling symptoms of Parkinson’s disease (PD) and is associated with balance impairment and increased falls risk, yet remains challenging to understand due to its episodic and context-dependent nature. Altered supplementary motor area (SMA) function at rest and before self-initiated movements has been implicated in FoG; however, whether this extends to functionally relevant balance behavior is unknown. A loss of balance in response to support surface translations during standing elicits a perturbation-evoked N1 potential, an error assessment signal linked to SMA activity. We therefore predicted that people with FoG would have smaller and later N1 responses than people with PD without FoG and older adults. We recorded electroencephalography (EEG) during backward support-surface translations in 29 people with PD (71 ± 7 years), including 13 people with a history of FoG (FoG+) and 16 people without FoG (FoG-), and 26 older adults (OA; 67 ± 6 years). N1 amplitude and latency were quantified at a central midline electrode and defined as the largest negative peak 100-300 ms post-perturbation. N1 responses were similar between OA and FoG-(Δ = 4.0 µV and 2.8 ms, respectively). In contrast, FoG+ had smaller N1 amplitudes than FoG- (Δ = 18.1 µV, p = 0.001) and OA (Δ = 14.1 µV, p = 0.003), and both differences persisted after adjusting for age. FoG+ also had later N1 latencies than OA (Δ = 26.5 ms, p = 0.029), with a similar but non-significant difference versus FoG- (Δ = 23.6 ms, p = 0.091); after adjusting for age, these latency differences were not retained.

These findings suggest that perturbation-evoked cortical activity is altered in FoG, potentially reflecting altered SMA function, and may provide a functionally relevant marker of FoG susceptibility.

## 1. INTRODUCTION

Freezing of gait (FoG) is one of the most disabling features of Parkinson’s disease (PD) and is associated with increased fall risk (Okuma et al., 2018; Perez-Lloret et al., 2014), yet how neural processes for mobility differ in individuals with and without FoG remains poorly understood. FoG can occur across disease stages and becomes increasingly prevalent with disease progression, affecting up to 80% of people with PD in advanced stages (Gilat et al., 2026; Gülke and Lewis, 2026). Although FoG manifests episodically, susceptibility to freezing may also reflect differences in neural processes that are evident even outside overt freezing episodes. For instance, even outside of freezing episodes, individuals with FoG exhibit poorer gait (Barbe et al., 2014; Glover et al., 2020), balance (Bekkers et al., 2017), and cognitive function (Monaghan et al., 2023) than those without a history of freezing, although the extent of these impairments varies across individuals. Identifying objective physiological markers of FoG susceptibility could improve our mechanistic understanding and inform the development of targeted assessments and interventions.

Dysfunction across widespread cortical and subcortical networks has been implicated in FoG, with many studies implicating the supplementary motor area (SMA) as an important node within these networks. The SMA is extensively connected with cortical and subcortical regions and contributes to internally generated movement, motor preparation, task switching, response inhibition, and balance control (de Lima-Pardini et al., 2020; Jacobs et al., 2009; Nachev et al., 2008; Rahimpour et al., 2022). Resting-state fMRI (Mi et al., 2017) and task-based fMRI using motor imagery (Peterson et al., 2014) or virtual-reality paradigms (Ehgoetz Martens et al., 2018; Gilat et al., 2018, 2015; Matar et al., 2019) have demonstrated altered SMA activity and functional connectivity with basal ganglia, premotor, and brainstem locomotor regions in people with FoG compared with those without FoG. However, the functional relevance of this network dysfunction remains unclear since fMRI is conducted in a supine, stationary scanner environment that does not challenge balance or pose a threat of falling. Functional near-infrared spectroscopy (fNIRS) has enabled measurement of cortical activity during actual locomotion, revealing altered recruitment of SMA and other frontal motor regions during FoG-provoking tasks such as turning and doorway passage, even in the absence of overt freezing episodes (Cockx et al., 2024). However, because hemodynamic signals are inherently slow, a limitation of both fMRI and fNIRS, the time course of altered SMA activity in balance and gait in people with FoG is unclear.

Standing balance perturbations elicit a prominent cortical negative potential (N1) measured by electroencephalography (EEG), with source-localization studies implicating the SMA as a primary contributor (Marlin et al., 2014; Mirdamadi et al., 2024; Protzak et al., 2026). The N1 is thought to reflect a balance error signal shaped by bottom-up and top-down factors (Payne et al., 2019; Purohit and Bhatt, 2022; Varghese et al., 2017). For example, N1 amplitudes scale with perturbation acceleration (Payne et al., 2019) and are modulated by perturbation predictability, attention, and postural threat (Adkin et al., 2008; Mochizuki et al., 2008; Quant et al., 2004). Perturbation-evoked N1 responses also demonstrate excellent within-individual reliability for at least one year in older adults (Mirdamadi et al., 2025), supporting its potential clinical utility as a biomarker in PD. In older adults, larger N1 responses correlate with poorer cognitive set-shifting (Payne et al., 2021), suggesting that individual differences in the N1 may relate to the ability to flexibly adapt balance responses to changing task demands. Cognitive set-shifting is also impaired in people with FoG (Monaghan et al., 2023), providing a potential link between N1-related cortical processing and freezing susceptibility.

Although our prior work found similar N1 responses between older adults and people with PD (Payne et al., 2022), whether N1 responses differ by freezing status remains unknown. Given the prominent contribution of SMA activity to the perturbation-evoked N1 (Protzak et al., 2026), the N1 may provide a physiologically relevant measure of altered cortical engagement during balance control in individuals with FoG, even in the absence of an overt freezing episode. Conceptual models propose that FoG arises when motor, cognitive, and limbic deficits accumulate or interfere with one another, limiting the ability to adapt ongoing movement to changes in context (Nieuwboer and Giladi, 2013). If FoG reflects a reduced capacity of SMA-implicated networks to process and respond to such changes, then people with FoG may exhibit altered perturbation-evoked cortical responses during balance control.

Here, we measured perturbation-evoked N1 responses using EEG during standing support-surface translations in people with PD with FoG (FoG+), people with PD without FoG (FoG-), and older adults. We predicted that FoG+ would have smaller N1 amplitudes than FoG- and older adults, reflecting reduced SMA engagement, and later N1 latencies, consistent with delayed N1 responses in populations with impaired balance (Duckrow, 1999; Palmer et al., 2025). As exploratory analyses, we examined whether N1 amplitude and latency were associated with freezing severity and other clinical characteristics relevant to FoG.

## 2. METHODS

### 2.1 Participants

The study included 29 people with PD (8 female, 70.8 ± 7.0 years old) and 26 older adults (OA; 14 female, 66.6 ± 6.5 years old). People with PD were eligible if they had a clinical diagnosis of PD, were taking dopaminergic medications, and were in Hoehn & Yahr Stage II-III. All participants with PD were Hoehn & Yahr Stage II, except for two participants who were Stage III (one FoG+, one FoG-). All participants were required to be 50-85 years of age, have no neurological conditions other than PD, have sufficient cognitive capacity to provide informed consent, be able to walk 10 m, and be able to stand for 10 minutes. The experimental protocol was approved by the Emory University Institutional Review Board, and all participants provided written informed consent. Data from a subset of OA (n = 14) were included in a previous study addressing different research questions (Mirdamadi et al., 2025).

### 2.2 Experimental Design

The present study included data from a single testing session in which people with PD were tested in the practically defined “OFF” state (>12 hours after dopaminergic medication intake) (McKay et al., 2019), except for two participants for whom data from the ON-medication session were used: one participant with FoG who was unable to tolerate OFF-medication testing and one participant without FoG who did not complete the OFF-medication session. These procedures were part of a larger project in which participants completed two sessions that differed in medication state (“OFF” and “ON” their typical dopaminergic medications), separated by approximately one week, with session order counterbalanced across participants; data from the ON-medication sessions will be reported separately. The present analyses were restricted to the OFF state, where available, to minimize the acute effects of dopaminergic medication and variability associated with medication dose and timing.

### 2.3 Freezing of Gait Assessment

FoG status was determined from clinical documentation and self-report. All participants with PD completed the New Freezing of Gait Questionnaire (NFOG-Q) (Nieuwboer et al., 2009) and item 3.11 of the MDS-UPDRS-III (Goetz et al., 2008). Participants also performed two consecutive 360° turns, one in each direction, to attempt to provoke FoG during the laboratory assessment. Participants were categorized as FoG+ (n = 13) if FoG was documented in their clinical record or self-reported on the NFOG-Q (total score > 0); all other participants with PD were classified as FoG- (n = 16). The NFOG-Q total score was used as a continuous measure of FoG severity in exploratory correlation analyses.

### 2.4 Clinical assessments and questionnaires

Clinical assessments were performed in the OFF-medication state, except for the Montreal Cognitive Assessment (MoCA) (Nasreddine et al., 2005), which was assessed in the ON-medication state and used as a measure of global cognitive function. Balance was assessed using the Mini-Balance Evaluation Systems Test (Mini-BESTest) (Leddy et al., 2011), and motor symptom severity was assessed using the MDS-UPDRS-III (Goetz et al., 2008). The MDS-UPDRS-III was video-recorded and scored offline by a neurologist, with the exception of rigidity, which was scored at the time of assessment. Cognitive set-shifting was assessed using the Trail Making Test (TMT) Parts A and B. Performance was quantified as the difference in completion time between Part B, which requires alternating between sequential numbers and letters, and Part A, which requires connecting numbers sequentially (TMT B-A). Larger values indicate poorer set-shifting ability (Sánchez-Cubillo et al., 2009). Balance confidence and anxiety, additional clinical characteristics relevant to FoG, were assessed using the Activities-specific Balance Confidence (ABC) Scale (Powell and Myers, 1995) and Beck Anxiety Inventory (BAI) (Beck et al., 1988), respectively.

### 2.5 Standing balance perturbations

Participants underwent a series of standing balance perturbations while EEG was continuously recorded (Section 2.6). Participants stood barefoot with their feet approximately hip-width apart and wore an overhead safety harness. Before testing, participants completed a familiarization block of backward perturbations to ensure that they could recover balance without taking a step.

Participants then underwent approximately 45 pairs of backward support-surface translation perturbations (7.5 cm, 15 cm/s, 0.01 m/s^2^) with their eyes closed. After each pair, participants verbally reported whether the second perturbation was in the same or a different direction from the first. In each pair, the first perturbation was directed directly backward and occurred at an unpredictable time. The second perturbation occurred 4 s later and was directed posterolateral to the left or right. Following the participant’s response, the platform returned to its initial position without lateral displacement to avoid providing additional directional information. Intertrial intervals ranged from approximately 10 to 60 s. Seated rest breaks were taken every 15 trials or sooner if requested. Additional details of the paradigm have been reported previously (Bong et al., 2020; Mirdamadi et al., 2025, 2024). Because the angle and direction of the second perturbation varied across trials and participants based on perceptual responses, only responses to the first perturbation were included in the present analyses.

### 2.6 EEG Recording and Preprocessing

EEG was continuously recording during standing balance perturbations (sampling frequency 1000Hz, impedance <25kOhm) using a 64-channel active electrode cap (actiCAP) and ActiCHamp amplifier with a 24-bit A/D converter and an online 20 kHz anti-aliasing low-pass filter (Brain Products, GmbH). Fz was the online reference electrode and Fpz was the ground electrode.

EEG preprocessing involved a modified version of the BeMoBIL pipeline (Klug et al., 2022) that incorporates removal of line noise (Klug and Kloosterman, 2022), removal and interpolation of bad channels, reconstruction of the online Fz reference after average re-referencing, high-pass filter with a cutoff of 1.5 Hz, and adaptive mixture component analysis algorithm (AMICA) (Palmer et al., 2008). Modifications to the original pipeline were similar to that described in our prior manuscripts involving independent component (IC) selection that used multiple approaches to distinguish brain from non-brain ICs (Mirdamadi et al., 2025; Protzak et al., 2026). We rejected ICs that were identified by ICLabel (Pion-Tonachini et al., 2019) as “eye”, “heart”, or “channel noise”, ICs with a residual variance > 0.15, ICs in which the linear slope of the power spectrum between 2-40 Hz was >0, and any additional ICs labeled as “bad channels” using the TESA toolbox (Rogasch et al., 2017). The remaining ICs were visually inspected based on their dipole location, scalp map, power spectrum, and power-power coupling from the Powpowcat toolbox (Thammasan and Miyakoshi, 2020) and removed if applicable. The final IC solution was projected to the continuous, unfiltered dataset (but otherwise preprocessed), since high-pass filters >1 Hz used to improve the AMICA decomposition can distort the signal (de Cheveigné and Nelken, 2019; Tanner et al., 2015). Channel-level data were high pass filtered with a cutoff of 0.5 Hz, low pass filtered with a cutoff of 25 Hz, re-referenced to the mastoids, epoched -2 to 2 seconds around each perturbation, and baseline removal -150 to -50 milliseconds.

### 2.7 N1 Data Analysis

For each participant, the channel-based time series data were averaged across single-trial epochs for the first perturbation of each pair. The N1 response was defined as the largest negative response between 100-300 msec post-perturbation based on prior work demonstrating delays in the evoked response in older adults (Duckrow, 1999; Mirdamadi et al., 2025; Palmer et al., 2025). If more than one local minimum was identified, we chose the most negative local minimum. If no local minimum was identified, we chose the most negative value. We quantified the N1 from 3 central midline electrodes (FCz, Cz, CPz) to account for individual differences and selected the electrode that generated the largest peak response for further analysis (Protzak et al., 2026).

### 2.8 Statistical Analyses

Age was compared between OA, FoG+, and FoG-using a one-way ANOVA; where significant, post-hoc pairwise comparisons were computed with Tukey correction. Sex was compared using Fisher’s exact test, with Cohen’s h as the effect size for the difference in proportions. For PD-specific variables (disease duration, UPDRS-III, MoCA, Mini-BESTest, ABC Scale, TMT B−A, and Beck Anxiety), Welch’s independent samples t-tests were used to compare FoG+ and FoG-only.

To test whether perturbation-evoked cortical responses differed across groups, separate one-way ANOVAs were conducted for N1 amplitude and latency, with Group (FoG+, FoG-, OA) as the between-subjects factor. Three pairwise comparisons were specified a priori (FoG+ vs. FoG−, FoG− vs. OA, and FoG+ vs. OA), with Tukey adjustment for multiple comparisons. Effect sizes are reported as partial eta-squared for omnibus tests and Cohen’s *d* for pairwise comparisons.

Exploratory Spearman rank correlations were used to examine associations between N1 amplitude and latency and clinical measures, given the small within-group samples and the positively skewed distribution of N1 amplitude within FoG+. Within FoG+, we examined associations with freezing severity (NFOG-Q), including Pearson partial correlations adjusting for age. Associations with Mini-BESTest, MoCA, TMT B-A, UPDRS-III, ABC Scale, and Beck Anxiety Inventory scores were examined separately within FoG+ and FoG-.

As post hoc covariate analyses, N1 group comparisons were repeated with age as a covariate and, within the PD sample, with age and disease duration as covariates. Covariate-adjusted models are reported in the Supplementary Results (Table S1). Statistical significance was set at alpha < 0.05.

## 3. RESULTS

### FoG+ had lower balance confidence and poorer clinical balance than FoG−, but similar disease duration, motor severity, and global cognition

Demographic and clinical characteristics are reported in Table 1 (group-level) and Table 2 (individual-level). Age differed across groups (F(2,52) = 4.06, p = 0.023), with FoG+ participants older than OA, while FoG-participants were similar in age to both groups. Age was therefore retained as a covariate in post hoc covariate models (Table S1). Sex distribution differed, with OA having more women compared to either PD group.

**Table 1.** Participant Demographic and Clinical Characteristics.

| Characteristic | FoG- (n = 16) | FoG+ (n = 13) | Statistic | p | Effect size |
|---|---|---|---|---|---|
| Age (years) | 68.9 ± 6.6 | 73.1 ± 7.1 | t(25.0) = -1.61 | .120 | -0.61 |
| Sex (F/M) | 6/10 | 2/11 | Fisher's exact | .238 | 0.51 |
| ABC Scale (% /100) | 89.8 ± 8.8 | 75.4 ± 16.2 | t(17.7) = 2.86 | .011 | 1.13 |
| TMT B-A (s) | 59.3 ± 61.4 | 64.2 ± 51.3 | t(27.0) = -0.24 | .816 | -0.09 |
| Mini-BESTest ( /28) | 23.7 ± 1.9 | 21.6 ± 3.1 | t(18.8) = 2.10 | .049 | 0.82 |
| Disease duration (years) | 4.8 ± 3.5 | 5.3 ± 3.5 | t(25.9) = -0.35 | .727 | -0.13 |
| UPDRS-III ( /132) | 31.8 ± 12.7 | 28.5 ± 9.6 | t(24.0) = 0.77 | .449 | 0.29 |
| MoCA ( /30) | 26.2 ± 2.9 | 24.9 ± 2.1 | t(26.7) = 1.41 | .171 | 0.51 |
| Beck Anxiety Inventory ( /63) | 9.2 ± 6.3 | 10.9 ± 5.8 | t(23.8) = -0.71 | .485 | -0.28 |

*Note. Values are mean ± SD unless otherwise stated. FoG- = Parkinson's disease without freezing of gait; FoG+ = Parkinson's disease with freezing of gait. Mini-BESTest = Mini Balance Evaluation Systems Test; UPDRS-III = Movement Disorder Society Unified Parkinson's Disease Rating Scale Part III (OFF-medication state); MoCA = Montreal Cognitive Assessment (ON-medication state only); ABC = Activities-specific Balance Confidence Scale; TMT B-A = Trail Making Test B minus A completion time. Statistical comparisons were made between FoG+ and FoG- only, using Welch's independent-samples t-tests with Cohen's d as the effect size; the t statistic and d are signed as FoG- minus FoG+. Sex was compared using Fisher's exact test, with Cohen's h reported as the effect size for the difference in proportions. Degrees of freedom vary across outcomes due to missing data (FoG-: UPDRS-III n = 14, Beck Anxiety Inventory n = 13; see Table 2).*

**Table 2.** Individual Participant Characteristics and N1 Measures.

| ID | Sex | Age | PD Duration (yrs) | NFOG-Q | MoCA | ABC (%) | Mini-BESTest | UPDRS-III | Beck Anxiety | N1 Amplitude (µV) | N1 Latency (ms) |
|---|---|---|---|---|---|---|---|---|---|---|---|
| ***FoG- (n = 16)*** |  |  |  |  |  |  |  |  |  |  |  |
| 1 | M | 71-75 | 2.0 | 0 | 30 | 62.5 | 22 | — | — | 31.2 | 182 |
| 2 | F | 61-65 | 2.0 | 0 | 28 | 93.1 | 27 | — | — | 23.1 | 190 |
| 3 | M | 51-55 | 13.0 | 0 | 30 | 85.6 | 23 | 41 | — | 24.9 | 176 |
| 4 | F | 66-70 | 4.4 | 0 | 25 | 91.9 | 22 | 24 | 8 | 60.4 | 178 |
| 5 | F | 61-65 | 5.6 | 0 | 26 | 85.6 | 24 | 16 | 16 | 48.1 | 192 |
| 6 | M | 71-75 | 1.0 | 0 | 29 | 98.8 | 25 | 25 | 6 | 35.4 | 208 |
| 7 | M | 71-75 | 3.9 | 0 | 28 | 98.1 | 23 | 57 | 6 | 41.6 | 168 |
| 8 | M | 76-80 | 2.9 | 0 | 24 | 97.5 | 24 | 37 | 5 | 41.0 | 198 |
| 9 | M | 71-75 | 8.1 | 0 | 24 | 88.1 | 20 | 45 | 21 | 12.3 | 230 |
| 10 | M | 71-75 | 10.6 | 0 | 28 | 95.0 | 25 | 41 | 2 | 40.2 | 210 |
| 11 | M | 66-70 | 3.3 | 0 | 26 | 91.3 | 25 | 26 | 2 | 33.8 | 198 |
| 12 | F | 71-75 | 7.7 | 0 | 27 | 93.1 | 23 | 33 | 3 | 22.7 | 166 |
| 13 | M | 66-70 | 6.3 | 0 | 27 | 91.9 | 26 | 41 | 8 | 23.2 | 214 |
| 14 | M | 61-65 | 2.4 | 0 | 27 | 93.1 | 26 | 9 | 10 | 22.1 | 240 |
| 15 | F | 66-70 | 4.0 | 0 | 19 | 91.2 | 22 | 28 | 15 | 23.5 | 226 |
| 16* | F | 66-70 | 0.2 | 0 | 22 | 79.4 | 22 | 22 | 18 | 25.5 | 210 |
| ***FoG+ (n = 13)*** |  |  |  |  |  |  |  |  |  |  |  |
| 17 | M | 71-75 | 9.4 | 14 | 22 | 73.8 | 21 | 21 | 16 | 5.2 | 292 |
| 18 | M | 76-80 | 3.4 | 4 | 23 | 67.5 | 22 | 19 | 5 | 6.3 | 228 |
| 19 | M | 81-85 | 2.4 | 12 | 26 | 54.4 | 25 | 25 | 6 | 7.7 | 258 |
| 20 | F | 71-75 | 11.4 | 16 | 27 | 79.4 | 20 | 18 | 10 | 16.6 | 230 |
| 21 | M | 81-85 | 3.3 | 0 | 24 | 79.4 | 19 | 27 | 6 | 11.3 | 230 |
| 22 | M | 71-75 | 2.6 | 5 | 23 | 93.8 | 20 | 30 | 7 | 22.5 | 186 |
| 23 | F | 71-75 | 1.7 | 6 | 24 | 47.5 | 19 | 16 | 11 | 3.6 | 182 |
| 24 | M | 76-80 | 6.7 | 0 | 26 | 87.5 | 24 | 30 | 8 | 10.6 | 208 |
| 25 | M | 76-80 | 3.0 | 10 | 23 | 54.4 | 21 | 48 | 19 | 10.6 | 274 |
| 26 | M | 56-60 | 1.0 | 3 | 24 | 97.5 | 25 | 30 | 4 | 43.7 | 212 |
| 27* | M | 66-70 | 10.0 | 0 | 29 | 93.8 | 26 | 31 | 11 | 9.4 | 188 |
| 28 | M | 71-75 | 6.0 | 0 | 25 | 68.8 | 24 | 30 | 22 | 8.7 | 232 |
| 29 | M | 61-65 | 8.0 | 13 | 28 | 82.5 | 15 | 45 | 17 | 22.1 | 176 |

*Note.* Values rounded to one decimal place where applicable; — = not assessed. PD Duration = years since diagnosis. NFOG-Q = New Freezing of Gait Questionnaire; a score of 0 denotes no self-reported freezing in the past month (four FoG+ participants scored 0 but had documented FoG in their charts; see Methods 2.3). MoCA = Montreal Cognitive Assessment (ON-medication state); ABC = Activities-specific Balance Confidence Scale; Mini-BESTest = Mini Balance Evaluation Systems Test; UPDRS-III = Movement Disorder Society Unified Parkinson's Disease Rating Scale Part III (OFF-medication state); Beck Anxiety = Beck Anxiety Inventory; FoG- = people without FoG; FoG+ = people with FoG (by NFOG-Q or clinical documentation). *\*Data from the ON-medication session.*

FoG+ and FoG-did not differ in disease duration, motor symptom severity (UPDRS-III), global cognition (MoCA), set-shifting (TMT B-A), or anxiety. FoG+ had lower balance confidence (ABC Scale) and poorer balance (Mini-BESTest) than FoG- (Table 1). All participants with PD were Hoehn and Yahr stage II, except for one participant in each group who was stage III.

Two participants were tested ON medication: one with FoG who was unable to tolerate the OFF-medication state (Table 2, ID 27) and one without FoG who did not complete the OFF-medication session (Table 2, ID 16).

### N1 responses were smaller in FoG+ compared to FoG- and OA

Perturbation-evoked N1 responses were smaller and had a less defined negative peak in FoG+ compared to FoG- and OA (Figure 1A; individual traces in Figure 1B). N1 amplitudes differed between groups (main effect of Group: F(2,52) = 8.78, p = 0.001, partial η² = 0.25), with smaller N1 amplitudes in FoG+ (13.7 ± 10.7 µV) than in both FoG- (31.8 ± 12.2 µV; p = 0.001, d = -1.57) and OA (27.8 ± 12.8 µV; p = 0.003, d = - 1.16). In contrast, N1 amplitudes were comparable between OA and FoG- (p = 0.562, d = -0.32).

**Figure 1.**
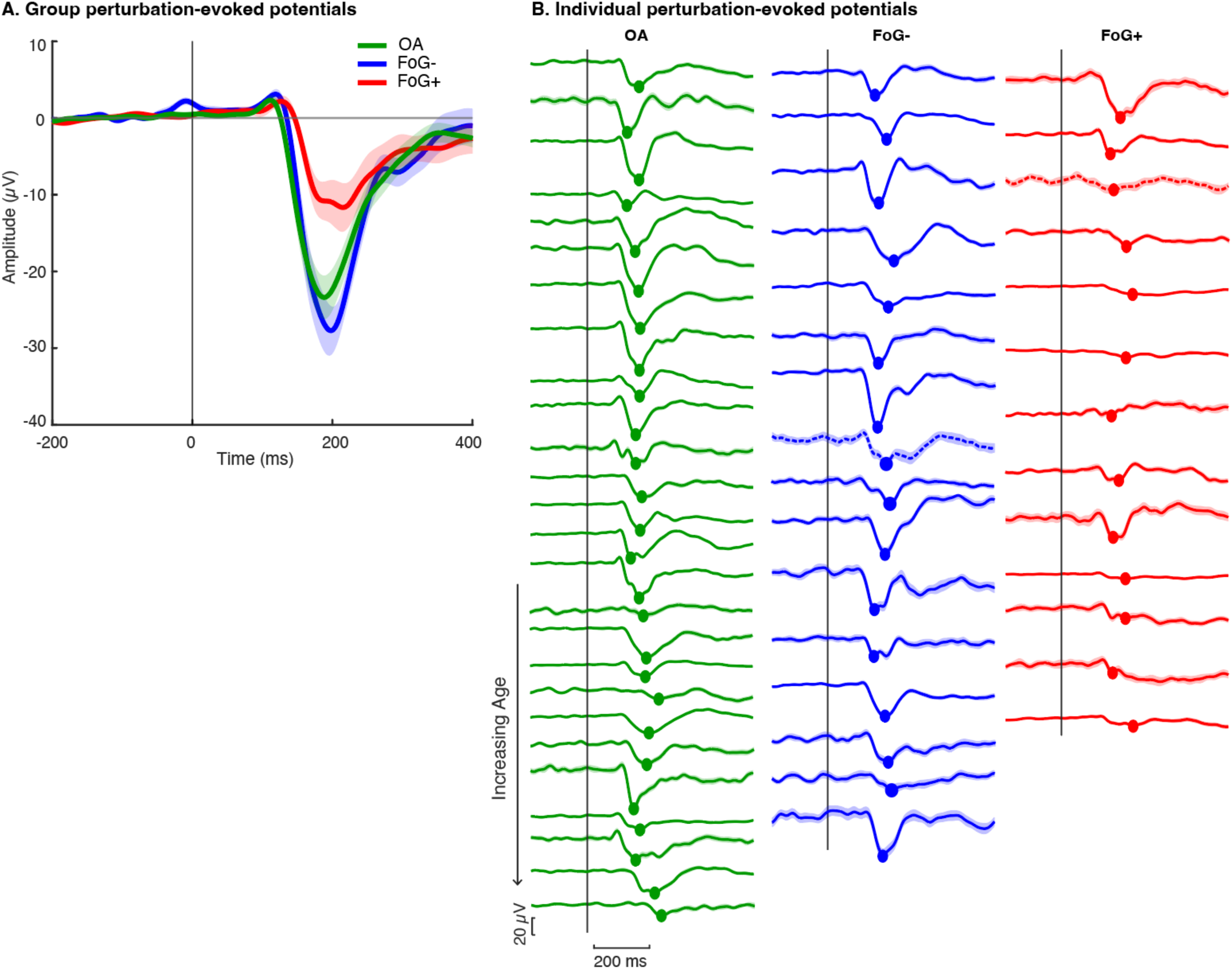
Perturbation-evoked cortical responses across groups and individuals. **A.** Grand average perturbation-evoked potentials for older adults (OA; green), people with Parkinson’s Disease without freezing of gait (FoG-; blue), and people with Parkinson’s Disease with freezing of gait (FoG+; red). Shaded region denotes standard error of the mean. Vertical line denotes perturbation onset. **B.** Individual EEG traces at the central electrode with the largest N1 peak response (100-300 ms post-perturbation), indicated by circles. Traces are ordered by age within each group (youngest to oldest). Dotted traces: participants with data collected in the ON-medication state. Vertical line denotes perturbation onset.

### N1 peak latencies were later in FoG+ compared to OA

N1 latencies differed between groups (main effect of Group: F(2,52) = 3.70, p = 0.031, partial η² = 0.13; Figure 2B). N1 latencies were comparable between OA (196 ± 30.0 ms) and FoG- (199 ± 22.2 ms; p = 0.952, d = 0.10). In contrast, N1 latency was later in FoG+ (223 ± 36.1 ms) compared to OA and tended to be later than in FoG- (p = 0.029, d = 0.82; p = 0.091, d = 0.81, respectively).

**Figure 2.**
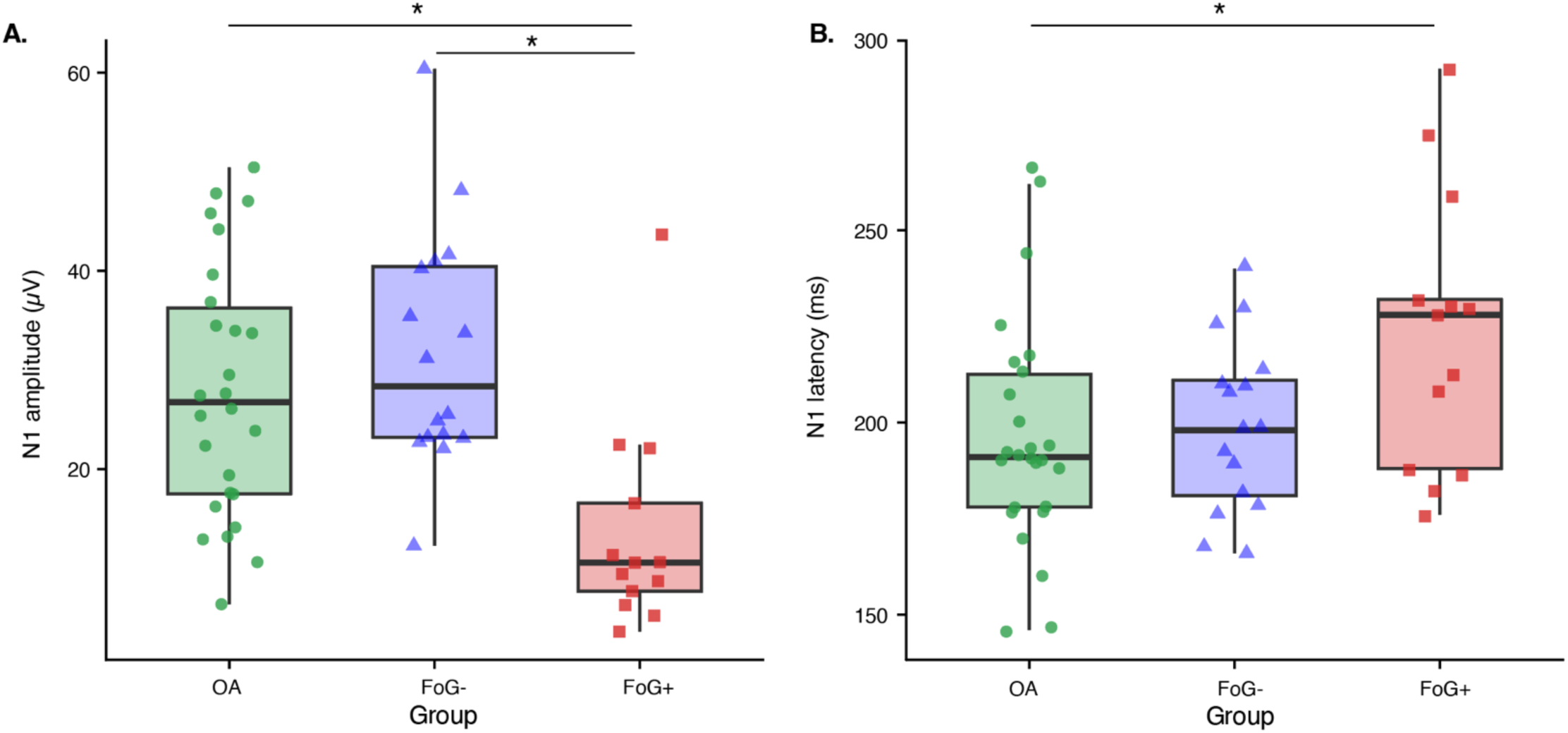
Group differences in N1 amplitude and latency. Group distributions of perturbation-evoked N1 amplitude (A) and peak latency (B) for OA (green), FoG- (blue), and FoG+ (red). Boxes represent the interquartile range; horizontal lines indicate the median. Significant pairwise differences (Tukey-corrected) are indicated by *; p < 0.05. OA = older adults without Parkinson’s Disease; FoG- = Parkinson’s disease without freezing of gait; FoG+ = Parkinson’s Disease with freezing of gait.

### Post-hoc Covariate Models

The primary group differences were re-examined post hoc with age as a covariate and, within the PD sample, with age and disease duration as covariates. Group differences in N1 amplitude were retained after adjusting for age (F(2,51) = 6.35, p = .003, partial η² = 0.20), with smaller N1 amplitudes in FoG+ than in both FoG- (p = .002) and OA (p = .039). Within the PD sample, the smaller N1 amplitude in FoG+ than in FoG- was also retained after adjusting for age and disease duration (β = -15.7 µV, p = .002). Group differences in latency were attenuated in both models (three-group: p = .28; PD only: p = .13). Age was a significant predictor of both N1 amplitude (β = -0.52 µV per year, p = .039) and N1 latency (β = 1.97 ms per year, p = .001). Full model results are reported in the Supplementary Results (Table S1).

### Exploratory associations between N1 responses and clinical outcomes

Within the FoG+ group, freezing severity (NFOG-Q) was not associated with N1 amplitude (ρ = -0.04, p = 0.89) or N1 latency (ρ = 0.20, p = 0.51; Supplementary Figure 1). Age-adjusted partial correlations were similarly non-significant (amplitude r = -0.07, p = 0.82; latency r = 0.46, p = 0.14).

N1 amplitude was not associated with balance performance, global cognition, set-shifting function, or anxiety in either group (all p > 0.20). Within FoG+, however, larger N1 amplitudes were associated with greater balance confidence (ABC Scale: ρ = 0.72, p = 0.006), which was not observed in FoG- (ρ = 0.17, p = 0.52) (Supplementary Figure 2). N1 amplitude was not significantly associated with motor symptom severity in either group (UPDRS-III: FoG+ ρ = 0.51, p = 0.075; FoG- ρ = -0.13, p = 0.65). N1 latency was not associated with any clinical outcomes in either group (all p > 0.08) (Supplementary Figure 3).

## 4. DISCUSSION

Here, we observed differences in perturbation-evoked cortical activity between people with PD with and without FoG during a functionally relevant balance task. Although the reactive balance task did not elicit overt freezing episodes, people with FoG demonstrated smaller N1 amplitudes and tended to have later N1 latencies than both people without FoG and older adults, with amplitude differences persisting after adjusting for age. In contrast, people with PD without FoG showed N1 responses similar to older adults, suggesting that altered cortical activity in PD may be more closely associated with FoG status than with PD diagnosis alone, and highlight the value of considering FoG status when examining cortical activity in PD. Importantly, these differences were evident even in the absence of a freezing episode, suggesting that perturbation-evoked cortical activity may capture neurophysiological differences associated with susceptibility to FoG.

FoG-related alterations in cortical activity may extend beyond voluntary movement to processes involved in responding to unexpected balance disturbances. Reduced cortical potentials associated with motor preparation have been observed in people with FoG compared to older adults during voluntary lower-limb movement and gait initiation (Karimi et al., 2021; Marquez et al., 2023). During balance perturbations, people with PD demonstrate altered preparatory cortical activity (Smith et al., 2012) but similar perturbation-evoked cortical potentials compared to older adults (Payne et al., 2022). However, neither study considered FoG status. Our findings extend prior work by identifying smaller N1 responses following unexpected balance disturbances in people with FoG, which may reflect altered cortical processes involved in rapidly evaluating and responding to unexpected balance disturbances. Consistent with this interpretation, both perturbation-evoked N1 responses and midfrontal theta activity (3-7 Hz), a marker of action monitoring (Luu et al., 2000), increase following a loss of balance, scaling with both the magnitude of the perturbation and need for corrective motor responses (Solis-Escalante et al., 2021). Importantly, midfrontal theta activity is reduced in people with FoG compared to those without FoG at rest and during lower-limb pedaling (Bosch et al., 2022; Singh et al., 2020), and reduced in people with PD with postural instability (Bosch et al., 2021), which frequently co-occurs with FoG (Bekkers et al., 2017). Together with smaller perturbation-evoked N1 potentials observed here, these findings suggest that people with FoG may have altered cortical processes associated with action monitoring that limits the ability to evaluate and response to balance perturbations.

Our findings support the hypothesis that FoG is associated with altered SMA and broader motor-network function, even without overt freezing episodes. In neurotypical adults, source localization analyses indicate that the perturbation-evoked balance N1 is dominated by a central midline cortical source encompassing the SMA (Protzak et al., 2026). Although source localization is beyond the scope of the present study, our findings of altered perturbation-evoked cortical activity in people with freezing are consistent with neuroimaging evidence implicating altered SMA activity and connectivity in FoG. Altered connectivity between the SMA and cortical and subcortical locomotor regions further suggests that FoG reflects dysfunction of a distributed motor network rather than the SMA in isolation (Bardakan et al., 2022; Fling et al., 2014; Song et al., 2021). One potentially relevant circuit is the hyperdirect pathway, through which the SMA and other frontal regions project directly to the subthalamic nucleus (STN) to rapidly modify or suppress motor output (Nambu et al., 2002). Altered cortical-STN signaling and recruitment of stopping-related circuitry have been implicated in FoG, particularly during challenging gait conditions such as turning and doorway passage (Cockx et al., 2024; Gilat et al., 2015; Matar et al., 2019). Thus, smaller N1 responses in FoG+ may reflect broader SMA-centered network dysfunction rather than isolated SMA dysfunction. Future studies combining EEG source localization with measures of cortical-subcortical connectivity could clarify the neural circuits underlying these altered responses.

Later perturbation-evoked N1 latencies may reflect slower cortical processing of balance-related information. This interpretation is consistent with the broader ERP literature, in which longer component latencies have been associated with slower stimulus evaluation and information processing (Pinal et al., 2015a; Terrasa et al., 2018), whereas ERP amplitude can reflect the magnitude of neural resource allocation or engagement during stimulus processing (Pinal et al., 2015b; Sachs et al., 2004). However, the latency differences observed here should be interpreted cautiously. Although N1 latency was later in FoG+ than in older adults, the difference between FoG+ and FoG-did not reach statistical significance. Moreover, group differences in N1 latency were attenuated after adjusting for age, which was itself a significant predictor of latency, and the FoG+ group was older than OA. Thus, later N1 responses may reflect age-related slowing of cortical processing rather than a FoG-specific alteration. Interpretation of N1 latency is further complicated because many individuals in the FoG+ group had atypical perturbation-evoked morphologies, with multiple small deflections often present within the N1 time window (100-300 ms post-perturbation) rather than a single well-defined peak. Thus, it is unclear whether latency estimates in FoG+ reflect true differences in response timing or are influenced by differences in overall waveform morphology.

Because people with FoG also demonstrated poorer balance, the smaller N1 responses observed in this group could reflect greater overall impairment rather than FoG status specifically. However, FoG+ and FoG-had similar disease duration, motor symptom severity, global cognition, and set-shifting. Prior work has associated N1 amplitude with individual differences in cognitive set-shifting and balance-related function in older adults (Payne et al., 2022). However, N1 amplitude was not associated with balance or set-shifting function within either PD group in the present study. One possible explanation is that the same perturbation magnitude represented different levels of balance challenge across individuals, which can influence cortical responses to perturbations (Kerr et al., 2026; Payne and Ting, 2020). Alternatively, N1 amplitude may capture only one of several neural processes contributing to individual differences in clinical function.

In contrast to the lack of association with clinical balance function, larger N1 amplitudes were associated with greater perceived balance confidence on the ABC Scale in people with FoG. This direction is opposite to that reported in neurotypical older adults, in whom larger N1 amplitudes were associated with lower balance confidence (Payne et al., 2022). This distinction may be clinically relevant because discordance between perceived and objective balance ability has been independently associated with fall history in people with PD, suggesting that perceived balance provides information not fully captured by performance-based measures (Longhurst et al., 2025a, 2025b). These findings suggest that the relationships between brain activity and clinical characteristics may differ by FoG status; however, given the small sample and exploratory nature of this analysis, these findings should be replicated before drawing mechanistic conclusions.

Despite group-level differences in N1 amplitude, individual variability suggests that perturbation-evoked cortical activity does not map simply onto binary FoG status. This variability is consistent with the clinical and pathophysiological heterogeneity of FoG and the episodic nature of its presentation. In this study, one individual in the FoG-group had an N1 amplitude more than 1.5 SD below the FoG-mean, within the range observed in FoG+, whereas one FoG+ participant had an N1 amplitude nearly 3 SD above the FoG+ mean, exceeding the FoG-mean. This variability may reflect heterogeneity in freezing phenotype (e.g., levodopa-responsive versus levodopa-unresponsive FoG), the presence or absence of postural instability, cognitive status, or diagnostic classification (Bekkers et al., 2017; McKay et al., 2019; Nonnekes et al., 2015). Larger and more diverse cohorts will be needed to determine whether perturbation-evoked cortical responses can distinguish established FoG from prodromal vulnerability to freezing, or whether they are better conceptualized as markers of a broader balance-cognitive phenotype associated with freezing risk.

Taken together, perturbation-evoked cortical activity may provide a sensitive and functionally relevant neurophysiological signal for identifying freezing-related balance dysfunction and warrants further study in larger cohorts. Given the N1 is elicited during a functionally relevant balance task that engages cognitive, affective, and sensorimotor processes, it may be capable of capturing distinct mechanisms recruited during freezing in real-world settings compared to stationary laboratory paradigms. This capability is particularly relevant given that freezing categorization varies with the measure used (Duysens and Smits-Engelsman, 2026), and subjective self-reports do not always align with episodes observed in the laboratory or clinic due to the transient and context-dependent nature of freezing (Gilat et al., 2026). Combined with the reliability and clinical feasibility of the balance N1 (Mirdamadi et al., 2025), this work lays the foundation for future studies to examine whether perturbation-evoked cortical activity can track disease progression, predict clinical outcomes, or change in response to therapeutic interventions.

## Ethics approval

The protocol was approved by the Emory University Institutional Review Board, and all participants provided written informed consent.

## Data availability

De-identified data supporting the findings of this study are available from the corresponding author upon reasonable request.

## Data Availability

Individual participant demographic, clinical and N1 data are provided in Table 2. All other data produced in the present study are available upon reasonable request to the authors.

## Acknowledgements

We would like to thank our research participants. Study data were collected and managed using Research Electronic Data Capture (REDCap) database hosted at Emory University.

---

**Supplementary Figure 1.**
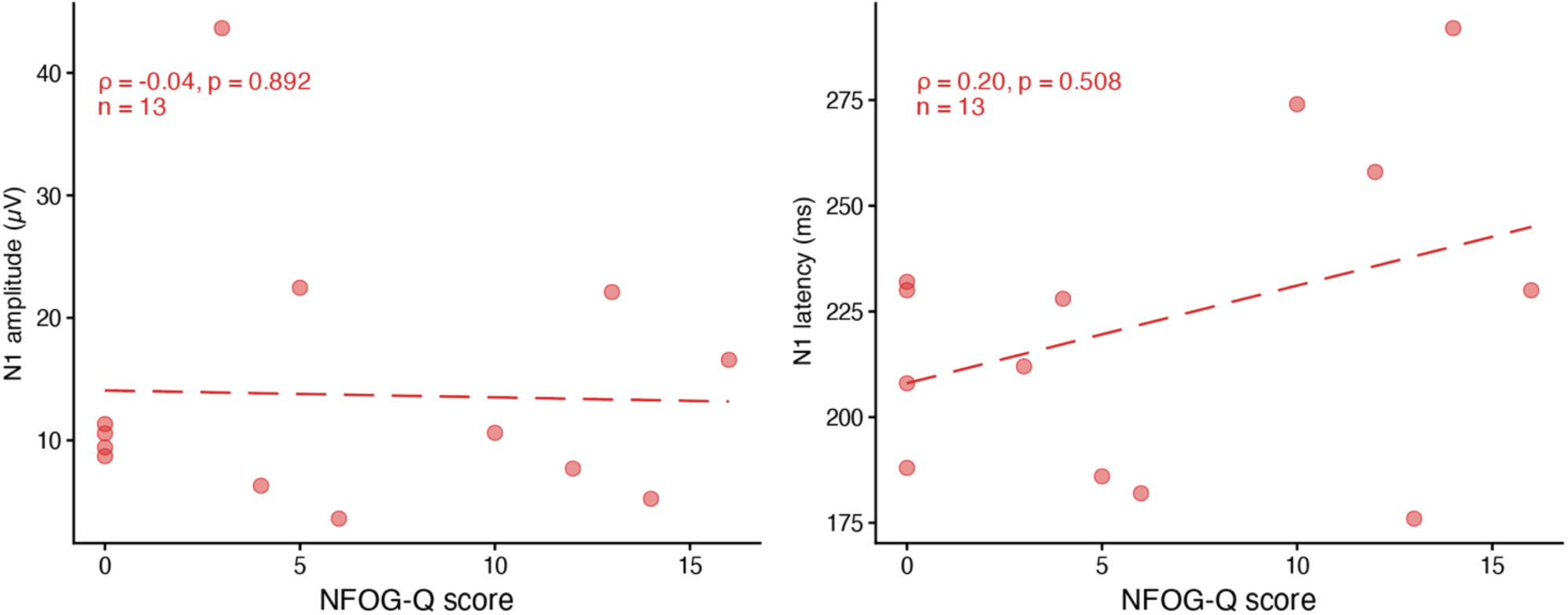
No association between NFOG-Q score and N1 peak amplitude or latency within the FoG+ group. Scatter plots showing the relationship between New Freezing of Gait Questionnaire (NFOG-Q) score and N1 peak amplitude and latency within FoG+ participants (n = 13). The regression line is shown. NFOG-Q scores of 0 reflect four participants whose FoG status was determined via clinical chart review or MDS-UPDRS item 3.11.

**Supplementary Figure 2.**
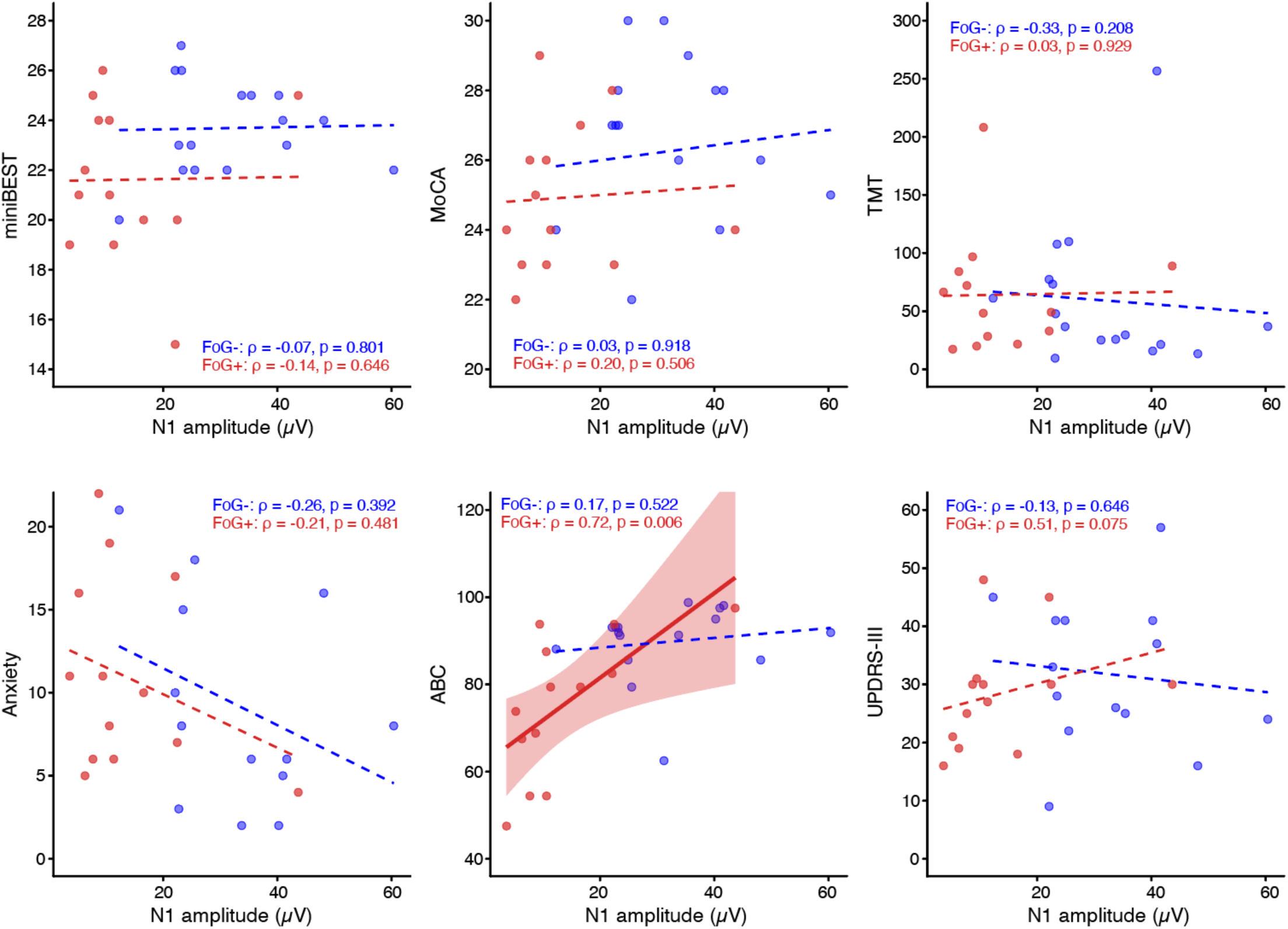
Within-group associations between N1 amplitude and clinical outcomes in FoG− and FoG+. Scatter plots showing Spearman correlations between N1 amplitude and each clinical outcome computed separately within FoG- (blue) and FoG+ (red). Regression lines are shown for each group. 95% confidence intervals are shown for significant correlations. Spearman’s ρ and p-value are annotated for each group within each panel. MoCA = Montreal Cognitive Assessment-MoCA was collected in the ON medication state; Anxiety was measured using the Beck Anxiety Inventory; TMT B-A = Trail Making Test B minus A completion time; ABC = Activities-specific Balance Confidence Scale; UPDRS-III = Movement Disorder Society Unified Parkinson’s Disease Rating Scale Part III.

**Supplementary Figure 3.**
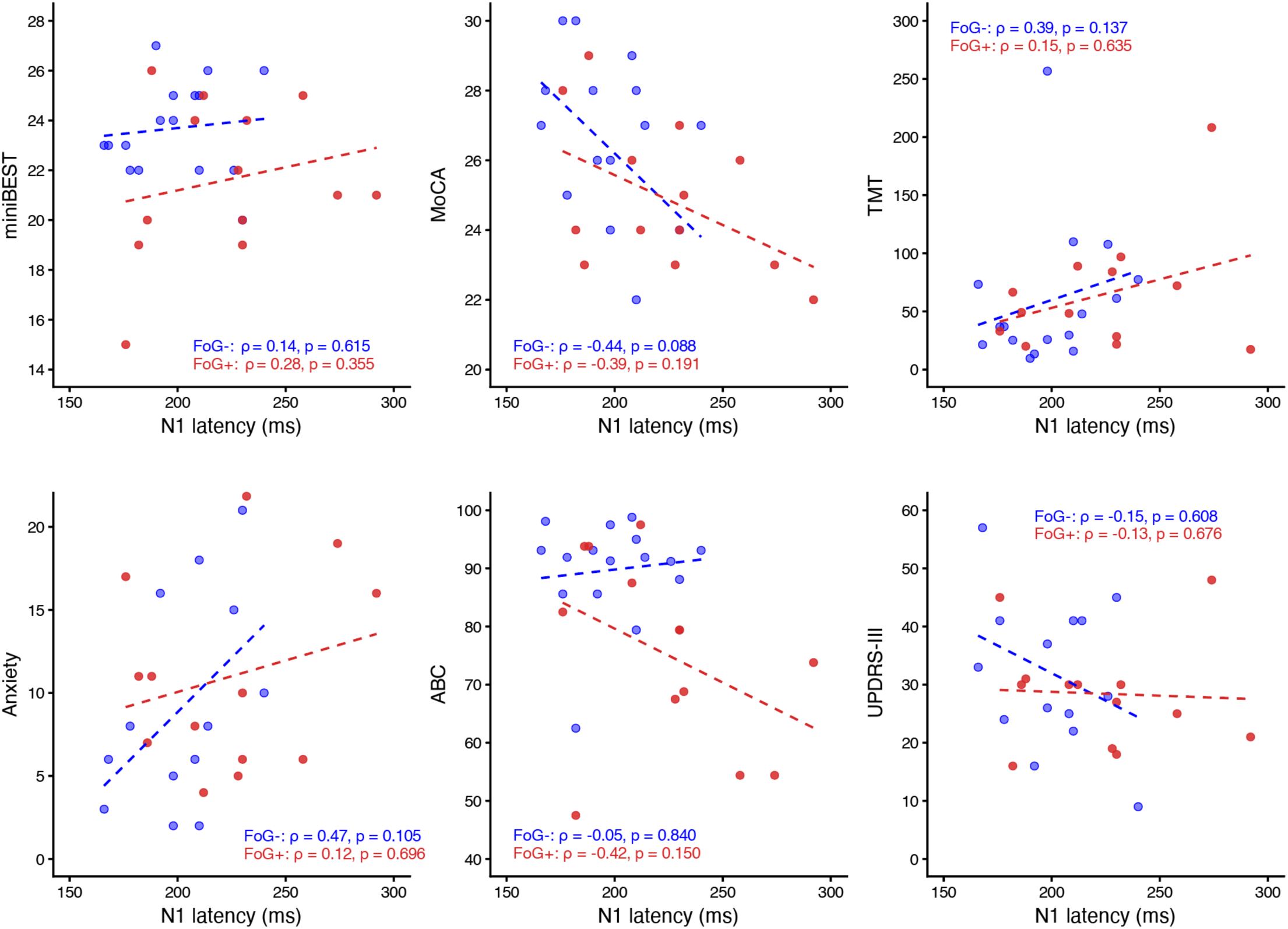
Within-group associations between N1 latency and clinical outcomes in FoG− and FoG+. Scatter plots showing Spearman correlations between N1 latency and each clinical outcome computed separately within FoG- (blue) and FoG+ (red). Regression lines are shown for each group. Spearman’s ρ and p-value are annotated for each group within each panel. MoCA = Montreal Cognitive Assessment-MoCA was collected in the ON medication state; TMT B-A = Trail Making Test B minus A completion time; Anxiety was measured using the Beck Anxiety Inventory. ABC = Activities-specific Balance Confidence Scale; UPDRS-III = Movement Disorder Society Unified Parkinson’s Disease Rating Scale Part III.

**Table S1.** Covariate-adjusted (post hoc) models for N1 amplitude and latency.

| Model | Effect / contrast | Statistic | p |
|---|---|---|---|
| A1.<br>Amplitude ~ age + group (N = 55) | Group \| age | F(2,51) = 6.35 | .003 |
|  | Age (covariate) | β = -0.52 | .039 |
|  | FoG- vs FoG+ (adj.) | — | .002 |
|  | FoG+ vs OA (adj.) | — | .039 |
| A2.<br>Latency ~ age + group (N = 55) | Group \| age | F(2,51) = 1.31 | .278 |
|  | Age (covariate) | β = 1.97 | .001 |
|  | FoG- vs FoG+ (adj.) | — | .294 |
|  | FoG+ vs OA (adj.) | — | .345 |
| B1. PD-only Amplitude ~ age + duration + group (n = 29) | FoG+ vs FoG- | β = -15.65 | .002 |
| B2. PD-only Latency ~ age + duration + group (n = 29) | FoG+ vs FoG- | β = 17.67 | .134 |

*Note. A1/A2, three-group ANCOVA with mean-centred age (N = 55); B1/B2, PD-only models with mean-centred age and median-centred disease duration (n = 29). “Group | age” is the omnibus effect of group adjusting for age. Adjusted pairwise contrasts are Tukey-corrected. β for age is per year; β for group is FoG+ relative to FoG-.*

## REFERENCES

1. Adkin, A.L., Campbell, A.D., Chua, R., Carpenter, M.G., 2008. The influence of postural threat on the cortical response to unpredictable and predictable postural perturbations. Neurosci. Lett. 435, 120–125. 10.1016/j.neulet.2008.02.018

2. Barbe, M.T., Amarell, M., Snijders, A.H., Florin, E., Quatuor, E.-L., Schönau, E., Fink, G.R., Bloem, B.R., Timmermann, L., 2014. Gait and upper limb variability in Parkinson’s disease patients with and without freezing of gait. J. Neurol. 261, 330–342. 10.1007/s00415-013-7199-1

3. Bardakan, M.M., Fink, G.R., Zapparoli, L., Bottini, G., Paulesu, E., Weiss, P.H., 2022. Imaging the neural underpinnings of freezing of gait in Parkinson’s disease. NeuroImage Clin. 35, 103123. 10.1016/j.nicl.2022.103123

4. Beck, A.T., Epstein, N., Brown, G., Steer, R.A., 1988. An inventory for measuring clinical anxiety: psychometric properties. J. Consult. Clin. Psychol. 56, 893–897. 10.1037//0022-006x.56.6.893

5. Bekkers, E.M.J., Dijkstra, B.W., Dockx, K., Heremans, E., Verschueren, S.M.P., Nieuwboer, A., 2017. Clinical balance scales indicate worse postural control in people with Parkinson’s disease who exhibit freezing of gait compared to those who do not: A meta-analysis. Gait Posture 56, 134–140. 10.1016/j.gaitpost.2017.05.009

6. Bong, S.M., McKay, J.L., Factor, S.A., Ting, L.H., 2020. Perception of whole-body motion during balance perturbations is impaired in Parkinson’s disease and is associated with balance impairment. Gait Posture 76, 44–50. 10.1016/j.gaitpost.2019.10.029

7. Bosch, T.J., Espinoza, A.I., Mancini, M., Horak, F.B., Singh, A., 2022. Functional Connectivity in Patients With Parkinson’s Disease and Freezing of Gait Using Resting-State EEG and Graph Theory. Neurorehabil. Neural Repair 36, 715–725. 10.1177/15459683221129282

8. Bosch, T.J., Kammermeier, S., Groth, C., Leedom, M., Hanson, E.K., Berg-Poppe, P., Singh, A., 2021. Cortical and Cerebellar Oscillatory Responses to Postural Instability in Parkinson’s Disease. Front. Neurol. 12. 10.3389/fneur.2021.752271

9. Cockx, H.M., Oostenveld, R., Flórez R, Y.A., Bloem, B.R., Cameron, I.G.M., van Wezel, R.J.A., 2024. Freezing of gait in Parkinson’s disease is related to imbalanced stopping–related cortical activity. Brain Commun. 6, fcae259. 10.1093/braincomms/fcae259

10. de Cheveigné, A., Nelken, I., 2019. Filters: When, Why, and How (Not) to Use Them. Neuron 102, 280–293. 10.1016/j.neuron.2019.02.039

11. de Lima-Pardini, A.C., Coelho, D.B., Nucci, M.P., Boffino, C.C., Batista, A.X., de Azevedo Neto, R.M., Silva-Batista, C., Barbosa, E.R., Cohen, R.G., Horak, F.B., Teixeira, L.A., Amaro Jr, E., 2020. Brain networks associated with anticipatory postural adjustments in Parkinson’s disease patients with freezing of gait. NeuroImage Clin. 28, 102461. 10.1016/j.nicl.2020.102461

12. Duckrow, R., 1999. Stance perturbation-evoked potentials in old people with poor gait and balance. Clin. Neurophysiol. 110, 2026–2032. 10.1016/S1388-2457(99)00195-9

13. Duysens, J., Smits-Engelsman, B., 2026. Freezing of Gait in Parkinson’s Disease: A Heterogeneous Phenomenon Across Assessment Tools. Eur. J. Neurosci. 63, e70572. 10.1111/ejn.70572

14. Ehgoetz Martens, K.A., Shine, J.M., Walton, C.C., Georgiades, M.J., Gilat, M., Hall, J.M., Muller, A.J., Szeto, J.Y.Y., Lewis, S.J.G., 2018. Evidence for subtypes of freezing of gait in Parkinson’s disease. Mov. Disord. Off. J. Mov. Disord. Soc. 33, 1174–1178. 10.1002/mds.27417

15. Fling, B.W., Cohen, R.G., Mancini, M., Carpenter, S.D., Fair, D.A., Nutt, J.G., Horak, F.B., 2014. Functional Reorganization of the Locomotor Network in Parkinson Patients with Freezing of Gait. PLOS ONE 9, e100291. 10.1371/journal.pone.0100291

16. Gilat, M., Ehgoetz Martens, K.A., Miranda-Domínguez, O., Arpan, I., Shine, J.M., Mancini, M., Fair, D.A., Lewis, S.J.G., Horak, F.B., 2018. Dysfunctional Limbic Circuitry Underlying Freezing of Gait in Parkinson’s Disease. Neuroscience 374, 119–132. 10.1016/j.neuroscience.2018.01.044

17. Gilat, M., Nonnekes, J., Factor, S.A., Bloem, B.R., Nutt, J.G., Giladi, N., Hallett, M., Nieuwboer, A., Horak, F.B., Weiss, D., Cubo, E., Zoetewei, D., Moreau, C., Jeon, B., Virmani, T., Hausdorff, J.M., Fasano, A., Lewis, S.J.G., 2026. An updated definition of freezing of gait. Nat. Rev. Neurol. 22, 172–181. 10.1038/s41582-025-01179-3

18. Gilat, M., Shine, J.M., Walton, C.C., O’Callaghan, C., Hall, J.M., Lewis, S.J.G., 2015. Brain activation underlying turning in Parkinson’s disease patients with and without freezing of gait: a virtual reality fMRI study. NPJ Park. Dis. 1, 15020. 10.1038/npjparkd.2015.20

19. Glover, A., Pillai, L., Doerhoff, S., Virmani, T., 2020. Differential Gait Decline in Parkinson’s Disease Enhances Discrimination of Gait Freezers from Non-freezers. J. Park. Dis. 10, 1657–1673. 10.3233/JPD-201961

20. Goetz, C.G., Tilley, B.C., Shaftman, S.R., Stebbins, G.T., Fahn, S., Martinez-Martin, P., Poewe, W., Sampaio, C., Stern, M.B., Dodel, R., Dubois, B., Holloway, R., Jankovic, J., Kulisevsky, J., Lang, A.E., Lees, A., Leurgans, S., LeWitt, P.A., Nyenhuis, D., Olanow, C.W., Rascol, O., Schrag, A., Teresi, J.A., Van Hilten, J.J., LaPelle, N., 2008. Movement Disorder Society-sponsored revision of the Unified Parkinson’s Disease Rating Scale (MDS-UPDRS): Scale presentation and clinimetric testing results. Mov. Disord. 23, 2129–2170. 10.1002/mds.22340

21. Gülke, E., Lewis, S.J.G., 2026. What is new in Freezing of Gait? J. Neurol. 273, 222. 10.1007/s00415-026-13765-6

22. Jacobs, J.V., Lou, J.S., Kraakevik, J.A., Horak, F.B., 2009. The supplementary motor area contributes to the timing of the anticipatory postural adjustment during step initiation in participants with and without Parkinson’s disease. Neuroscience 164, 877–885. 10.1016/j.neuroscience.2009.08.002

23. Karimi, F., Niu, J., Gouweleeuw, K., Almeida, Q., Jiang, N., 2021. Movement-related EEG signatures associated with freezing of gait in Parkinson’s disease: an integrative analysis. Brain Commun. 3, fcab277. 10.1093/braincomms/fcab277

24. Kerr, K.G., Boebinger, S.E., Mirdamadi, J.L., Protzak, J., Borich, M.R., Ting, L.H., 2026. Cortical response to balance perturbation is more sensitive in modern dancers than nondancers during biomechanically similar balance recovery. J. Neurophysiol. 10.1152/jn.00606.2025

25. Klug, M., Jeung, S., Wunderlich, A., Gehrke, L., Protzak, J., Djebbara, Z., Argubi-Wollesen, A., Wollesen, B., Gramann, K., 2022. The BeMoBIL Pipeline for automated analyses of multimodal mobile brain and body imaging data. 10.1101/2022.09.29.510051

26. Klug, M., Kloosterman, N.A., 2022. Zapline-plus: A Zapline extension for automatic and adaptive removal of frequency-specific noise artifacts in M/EEG. Hum. Brain Mapp. 43, 2743–2758. 10.1002/hbm.25832

27. Leddy, A.L., Crowner, B.E., Earhart, G.M., 2011. Utility of the Mini-BESTest, BESTest, and BESTest sections for balance assessments in individuals with Parkinson disease. J Neurol Phys Ther 35, 90–7. 10.1097/NPT.0b013e31821a620c

28. Longhurst, J.K., Hooyman, A., Albrecht, F., Franzén, E., Peterson, D.S., 2025a. Discordance Between Balance Ability and Perception and Its Relation to Falls in Parkinson’s Disease: A Replication Analysis. Neurorehabil. Neural Repair 39, 555–558. 10.1177/15459683251335316

29. Longhurst, J.K., Hooyman, A., Landers, M.R., Mancini, M., Franzén, E., Leavy, B., Johansson, H., Peterson, D., 2025b. Discordance Between Balance Ability and Perception Is Associated With Falls in Parkinson’s Disease: A Coordinated Analysis. Neurorehabil. Neural Repair 39, 114–125. 10.1177/15459683241300456

30. Luu, P., Flaisch, T., Tucker, D.M., 2000. Medial Frontal Cortex in Action Monitoring. J. Neurosci. 20, 464–469. 10.1523/JNEUROSCI.20-01-00464.2000

31. Marlin, A., Mochizuki, G., Staines, W.R., McIlroy, W.E., 2014. Localizing evoked cortical activity associated with balance reactions: does the anterior cingulate play a role? J. Neurophysiol. 111, 2634–2643. 10.1152/jn.00511.2013

32. Marquez, J.S., Bartsch, R.P., Günther, M., Hasan, S.M.S., Koren, O., Plotnik, M., Bai, O., 2023. Supplementary Motor Area Activity Differs in Parkinson’s Disease with and without Freezing of Gait. Park. Dis. 2023, 5033835. 10.1155/2023/5033835

33. Matar, E., Shine, J.M., Gilat, M., Ehgoetz Martens, K.A., Ward, P.B., Frank, M.J., Moustafa, A.A., Naismith, S.L., Lewis, S.J.G., 2019. Identifying the neural correlates of doorway freezing in Parkinson’s disease. Hum. Brain Mapp. 40, 2055–2064. 10.1002/hbm.24506

34. McKay, J.L., Goldstein, F.C., Sommerfeld, B., Bernhard, D., Perez Parra, S., Factor, S.A., 2019. Freezing of Gait can persist after an acute levodopa challenge in Parkinson’s disease. NPJ Park. Dis. 5, 25. 10.1038/s41531-019-0099-z

35. Mi, T.-M., Mei, S.-S., Liang, P.-P., Gao, L.-L., Li, K.-C., Wu, T., Chan, P., 2017. Altered resting-state brain activity in Parkinson’s disease patients with freezing of gait. Sci. Rep. 7, 16711. 10.1038/s41598-017-16922-0

36. Mirdamadi, J.L., Poorman, A., Munter, G., Jones, K., Ting, L.H., Borich, M.R., Payne, A.M., 2025. Excellent test-retest reliability of perturbation-evoked cortical responses supports feasibility of the balance N1 as a clinical biomarker. J. Neurophysiol. jn.00583.2024. 10.1152/jn.00583.2024

37. Mirdamadi, J.L., Ting, L.H., Borich, M.R., 2024. Distinct Cortical Correlates of Perception and Motor Function in Balance Control. J. Neurosci. Off. J. Soc. Neurosci. 44, e1520232024. 10.1523/JNEUROSCI.1520-23.2024

38. Mochizuki, G., Sibley, K.M., Esposito, J.G., Camilleri, J.M., McIlroy, W.E., 2008. Cortical responses associated with the preparation and reaction to full-body perturbations to upright stability. Clin. Neurophysiol. Off. J. Int. Fed. Clin. Neurophysiol. 119, 1626–1637. 10.1016/j.clinph.2008.03.020

39. Monaghan, A.S., Gordon, E., Graham, L., Hughes, E., Peterson, D.S., Morris, R., 2023. Cognition and freezing of gait in Parkinson’s disease: A systematic review and meta-analysis. Neurosci. Biobehav. Rev. 147, 105068. 10.1016/j.neubiorev.2023.105068

40. Nachev, P., Kennard, C., Husain, M., 2008. Functional role of the supplementary and pre-supplementary motor areas. Nat. Rev. Neurosci. 9, 856–869. 10.1038/nrn2478

41. Nambu, A., Tokuno, H., Takada, M., 2002. Functional significance of the cortico-subthalamo-pallidal “hyperdirect” pathway. Neurosci. Res. 43, 111–117. 10.1016/s0168-0102(02)00027-5

42. Nasreddine, Z.S., Phillips, N.A., Bédirian, V., Charbonneau, S., Whitehead, V., Collin, I., Cummings, J.L., Chertkow, H., 2005. The Montreal Cognitive Assessment, MoCA: A Brief Screening Tool For Mild Cognitive Impairment. J. Am. Geriatr. Soc. 53, 695–699. 10.1111/j.1532-5415.2005.53221.x

43. Nieuwboer, A., Giladi, N., 2013. Characterizing freezing of gait in Parkinson’s disease: Models of an episodic phenomenon. Mov. Disord. 28, 1509–1519. 10.1002/mds.25683

44. Nieuwboer, A., Rochester, L., Herman, T., Vandenberghe, W., Emil, G.E., Thomaes, T., Giladi, N., 2009. Reliability of the new freezing of gait questionnaire: Agreement between patients with Parkinson’s disease and their carers. Gait Posture 30, 459–463. 10.1016/j.gaitpost.2009.07.108

45. Nonnekes, J., Snijders, A.H., Nutt, J.G., Deuschl, G., Giladi, N., Bloem, B.R., 2015. Freezing of gait: a practical approach to management. Lancet Neurol. 14, 768–778. 10.1016/S1474-4422(15)00041-1

46. Okuma, Y., Silva de Lima, A.L., Fukae, J., Bloem, B.R., Snijders, A.H., 2018. A prospective study of falls in relation to freezing of gait and response fluctuations in Parkinson’s disease. Parkinsonism Relat. Disord. 46, 30–35. 10.1016/j.parkreldis.2017.10.013

47. Palmer, J.A., Makeig, S., Kreutz-Delgado, K., Rao, B.D., 2008. Newton method for the ICA mixture model, in: 2008 IEEE International Conference on Acoustics, Speech and Signal Processing. Presented at the ICASSP 2008. IEEE International Conference on Acoustic, Speech and Signal Processes, IEEE, Las Vegas, NV, pp. 1805–1808. 10.1109/ICASSP.2008.4517982

48. Palmer, J.A., Payne, A.M., Mirdamadi, J.L., Ting, L.H., Borich, M.R., 2025. Delayed Cortical Responses During Reactive Balance After Stroke Associated With Slower Kinetics and Clinical Balance Dysfunction. Neurorehabil. Neural Repair 39, 16–30. 10.1177/15459683241282786

49. Payne, A.M., McKay, J.L., Ting, L.H., 2022. The cortical N1 response to balance perturbation is associated with balance and cognitive function in different ways between older adults with and without Parkinson’s disease. Cereb. Cortex Commun. 3, tgac030. 10.1093/texcom/tgac030

50. Payne, A.M., Palmer, J.A., McKay, J.L., Ting, L.H., 2021. Lower Cognitive Set Shifting Ability Is Associated With Stiffer Balance Recovery Behavior and Larger Perturbation-Evoked Cortical Responses in Older Adults. Front. Aging Neurosci. 13, 742243. 10.3389/fnagi.2021.742243

51. Payne, A.M., Ting, L.H., 2020. Worse balance is associated with larger perturbation-evoked cortical responses in healthy young adults. Gait Posture 80, 324–330. 10.1016/j.gaitpost.2020.06.018

52. Payne, A.M., Ting, L.H., Hajcak, G., 2019. Do sensorimotor perturbations to standing balance elicit an error-related negativity? Psychophysiology 56, e13359. 10.1111/psyp.13359

53. Perez-Lloret, S., Negre-Pages, L., Damier, P., Delval, A., Derkinderen, P., Destée, A., Meissner, W.G., Schelosky, L., Tison, F., Rascol, O., 2014. Prevalence, Determinants, and Effect on Quality of Life of Freezing of Gait in Parkinson Disease. JAMA Neurol. 71, 884–890. 10.1001/jamaneurol.2014.753

54. Peterson, D.S., Pickett, K.A., Duncan, R.P., Perlmutter, J.S., Earhart, G.M., 2014. Brain activity during complex imagined gait tasks in Parkinson disease. Clin. Neurophysiol. Off. J. Int. Fed. Clin. Neurophysiol. 125, 995–1005. 10.1016/j.clinph.2013.10.008

55. Pinal, D., Zurrón, M., Díaz, F., 2015a. An Event Related Potentials Study of the Effects of Age, Load and Maintenance Duration on Working Memory Recognition. PLOS ONE 10, e0143117. 10.1371/journal.pone.0143117

56. Pinal, D., Zurrón, M., Díaz, F., 2015b. Age-related changes in brain activity are specific for high order cognitive processes during successful encoding of information in working memory. Front. Aging Neurosci. 7. 10.3389/fnagi.2015.00075

57. Pion-Tonachini, L., Kreutz-Delgado, K., Makeig, S., 2019. ICLabel: An automated electroencephalographic independent component classifier, dataset, and website. NeuroImage 198, 181–197. 10.1016/j.neuroimage.2019.05.026

58. Powell, L.E., Myers, A.M., 1995. The Activities-specific Balance Confidence (ABC) Scale. J. Gerontol. A. Biol. Sci. Med. Sci. 50A, M28–34. 10.1093/gerona/50a.1.m28

59. Protzak, J., Mirdamadi, J.L., Borich, M.R., Ting, L.H., 2026. A primary central source determines perturbation-evoked N1 amplitudes in younger adults. J. Neurophysiol. 135, 1315–1323. 10.1152/jn.00002.2026

60. Purohit, R., Bhatt, T., 2022. Mobile Brain Imaging to Examine Task-Related Cortical Correlates of Reactive Balance: A Systematic Review. Brain Sci. 12, 1487. 10.3390/brainsci12111487

61. Quant, S., Adkin, A.L., Staines, W.R., Maki, B.E., McIlroy, W.E., 2004. The effect of a concurrent cognitive task on cortical potentials evoked by unpredictable balance perturbations. BMC Neurosci. 5, 18. 10.1186/1471-2202-5-18

62. Rahimpour, S., Rajkumar, S., Hallett, M., 2022. The Supplementary Motor Complex in Parkinson’s Disease. J. Mov. Disord. 15, 21–32. 10.14802/jmd.21075

63. Rogasch, N.C., Sullivan, C., Thomson, R.H., Rose, N.S., Bailey, N.W., Fitzgerald, P.B., Farzan, F., Hernandez-Pavon, J.C., 2017. Analysing concurrent transcranial magnetic stimulation and electroencephalographic data: A review and introduction to the open-source TESA software. NeuroImage 147, 934–951. 10.1016/j.neuroimage.2016.10.031

64. Sachs, G., Anderer, P., Margreiter, N., Semlitsch, H., Saletu, B., Katschnig, H., 2004. P300 event-related potentials and cognitive function in social phobia. Psychiatry Res. 131, 249–261. 10.1016/j.pscychresns.2004.05.005

65. Sánchez-Cubillo, I., Periáñez, J.A., Adrover-Roig, D., Rodríguez-Sánchez, J.M., Ríos-Lago, M., Tirapu, J., Barceló, F., 2009. Construct validity of the Trail Making Test: role of task-switching, working memory, inhibition/interference control, and visuomotor abilities. J. Int. Neuropsychol. Soc. JINS 15, 438–450. 10.1017/S1355617709090626

66. Singh, A., Cole, R.C., Espinoza, A.I., Brown, D., Cavanagh, J.F., Narayanan, N.S., 2020. Frontal theta and beta oscillations during lower-limb movement in Parkinson’s disease. Clin. Neurophysiol. 131, 694–702. 10.1016/j.clinph.2019.12.399

67. Smith, B.A., Jacobs, J.V., Horak, F.B., 2012. Effects of magnitude and magnitude predictability of postural perturbations on preparatory cortical activity in older adults with and without Parkinson’s disease. Exp. Brain Res. 222, 455–470. 10.1007/s00221-012-3232-3

68. Solis-Escalante, T., Stokkermans, M., Cohen, M.X., Weerdesteyn, V., 2021. Cortical responses to whole-body balance perturbations index perturbation magnitude and predict reactive stepping behavior. Eur. J. Neurosci. 54, 8120–8138. 10.1111/ejn.14972

69. Song, W., Raza, H.K., Lu, L., Zhang, Z., Zu, J., Zhang, W., Dong, L., Xu, C., Gong, X., Lv, B., Cui, G., 2021. Functional MRI in Parkinson’s disease with freezing of gait: a systematic review of the literature. Neurol. Sci. 42, 1759–1771. 10.1007/s10072-021-05121-5

70. Tanner, D., Morgan-Short, K., Luck, S.J., 2015. How inappropriate high-pass filters can produce artifactual effects and incorrect conclusions in ERP studies of language and cognition. Psychophysiology 52, 997– 1009. 10.1111/psyp.12437

71. Terrasa, J.L., Montoya, P., González-Roldán, A.M., Sitges, C., 2018. Inhibitory Control Impairment on Somatosensory Gating Due to Aging: An Event-Related Potential Study. Front. Hum. Neurosci. 12. 10.3389/fnhum.2018.00280

72. Thammasan, N., Miyakoshi, M., 2020. Cross-Frequency Power-Power Coupling Analysis: A Useful Cross-Frequency Measure to Classify ICA-Decomposed EEG. Sensors 20, 7040. 10.3390/s20247040

73. Varghese, J.P., McIlroy, R.E., Barnett-Cowan, M., 2017. Perturbation-evoked potentials: Significance and application in balance control research. Neurosci. Biobehav. Rev. 83, 267–280. 10.1016/j.neubiorev.2017.10.022

